# Monitoring of azathioprine metabolite concentrations and cytokine levels in neuromyelitis optica spectrum disorder

**DOI:** 10.1101/2023.06.08.23290121

**Authors:** Qingmeng Huang, Junjie Wei, Chunfang Luo, Junhui Qin, Chunnv Tang, Lijing Li, Huijie Zhou, Kang Zhong, Bailing Lin, Yulan Tang

**Author notes:** **Correspondence authors** Yulan Tang, Department of Neurology, The First Affiliated Hospital of Guangxi Medical University, Nanning, 530021,China.

## Abstract

**Background:** The pathogenesis of NMOSD has been linked to the cytokines interleukins (IL) -6, NOD-, LRR-and pyrin domain-containing 3 (NLRP3) and IL-18 that contribute to development of inflammatory reactionsmay. Although azathioprine (AZA) is efficacious in preventing NMOSD recurrence, it may have adverse effects (AEs) maybe related to the plasma concentrations.

**Objective:** We would monitor the blood concentrations of azathioprine in NMOSD, and their relationship with cytokines, severity, efficacy, and safety range of the drug.

**Methods:** A total of 53 NMOSD patients were included in the study, which included 20 patients who had received AZA treatment within one month, and 16 patients who had received AZA treatment within six months, as well as 17 patients who had received AZA treatment over one year. The patient’s immunotherapy regimen was low-dose hormone combined with AZA. AZA was started at small doses and added every two weeks after no AEs, namely 50 mg qd for two weeks, 50mg bid for two weeks, and maintained at 50mg tid. The following clinical data were collected: gender, age, clinical symptoms, EDSS score, number of recurrences and AEs, etc. Healthy controls (HC) comprised 10 individuals. AZA metabolite concentrations 6-thioguaninenucleotides (6-TGN) and 6-methylmercaptopurine nucleotides (6-MMPN) were measured by High-performance liquid chromatography (HPLC). Levels of IL-6, NLRP3 and IL-18 were measured by Enzyme-linked immunosorbent assay (ELISA).

**Results:** Treatment with AZA decreased the EDSS score and the ARR. The 6-TGN and 6-MMPN concentrations gradually increased as the medication time increased, and was basically stable by 6 months of medication. EDSS score improvement was positively correlated with 6-TGN concentrations. Patients with AEs had higher concentrations of 6-TGN and 6-MMPN than those without. The serum levels of IL-6, NLRP3, and IL-18 were higher in the patients than in the HC. NLRP3 level was higher in neuropathic pain (NP) patients than in those without NP.

**Conclusion:** Consequently, AZA was confirmed to be an effective drug used to treat NMOSD. AEs increased when 6-TGN reached 155.604 pmol/8×10^8^ RBC or 6-MMPN reached 2583.168 pmol/8×10^8^ RBC. In NMOSD remission, IL-6 and NLRP3 levels were significantly higher than in normal subjects, suggesting that they may contribute to the pathogenesis of NMOSD. A high level of NLRP 3 may lead to NP.

## 1. Introduction

Neuromyelitis optica spectrum disorder (NMOSD) is an autoimmune-mediated demyelinating disorder of the central nervous system (CNS) which often causes severe disability and even death.. There has been evidence that AZA lowers EDSS scores and reduces ARR in NMOSD patients ^[1, 2]^. AZA mainly undergoes competitive metabolism through the following three pathways: a. A thiopurine methyl transferase (TPMT) catalyzes the formation of the inactive intermediate metabolite 6-MMPN. b. Xanthine oxidase generates the final metabolite, thiuric acid, which is excreted through the kidneys. c. Hypoxanthine phosphoribosyltransferase generates 6-TGN through the action of enzymes, and acts through the infiltration of DNA and RNA into lymphocytes, resulting in nonfunctional nucleic acids, inhibiting DNA, RNA, and protein synthesis, thereby exerting a cytotoxic effect, and preventing lymphocyte proliferation. The concentration of 6-TGN and 6-MMPN could be detected by High-performance liquid chromatography (HPLC)^[3]^.AZA treatment may also have the related adverse reactions (AEs), such as leukopenia, abnormal liver function, gastrointestinal symptoms, alopecia, and a menstrual disorder for women. In addition to evaluating the drug efficacy and predicting AEs, drug concentration monitoring can also be used to help adjust individual treatment doses.

Aquaporin-4 immunoglobulin G antibodies (AQP4-IgG) combine with AQP-4 of astrocyte foot processes in NMOSD, then leading to neuroinflammation^[4]^. As a proinflammatory cytokine, IL-6 stimulates antibody production and the development of effector T-cells in the immune response, Interfering with the function of the BBB and regulating the balance between regulatory and helper T-cells^[5]^. Plasma cells were induced to differentiate into plasma cells by IL-6, and pathogenic AQP-4 IgG was secreted by plasma cells in response to IL-6^[6]^. NLRP3 inflammasome is a key component of the innate immune system,wihch is mainly expressed in immune cells, also expressed in microglia and neurons in CNS^[7]^.It mediates caspase-1 activation and the pro-inflammatory cytokine IL-1 β, IL-18, in response to microbial infections and cell damage. Abnormal activation of the NLRP3 inflammasome is associated with inflammatory diseases, and its main role is to promote secretion of inflammatory cytokines and induction of pyroptosis^[8]^. The CSF NLRP3 level is increased in NMOSD patients, that may be related to mitochondrial dysfunction^[9]^.IL-18 is involved in the regulation of innate and acquired immune responses. IL-18 also is associated with the pathogenesis of immune diseases^[10, 11]^. It has been shown that higher blood levels of IL-18 and caspase-1 mRNA are associated with severity of MS^[12, 13]^.

In this study, we detected the concentration of AZA metabolites 6-TGN and 6-MMN in NMOSD patients by HPLC. We investigated the changes in drug concentrations after AZA treatment, and the correlation between drug concentrations and clinical indicators. The serum IL-6, NLRP3 inflammasome and IL-18 levels in NMOSD remission are detected used Enzyme-linked immunosorbent assay (ELISA), in order to understand the pathogenesis of NMOSD.

## 2. Materials and methods

### 2.1. Patients and controls

A total of 53 NMOSD patients were recruited from Neurology Department of the First Affiliated Hospital of Guangxi Medical University. All patients with NMOSD met the international consensus diagnostic criteria for NMOSD published in 2015^[14]^. Before medication, TPMT activity was normal, and AZA was started at small doses and added every two weeks after no AEs, namely 50 mg qd for two weeks, 50mg bid for two weeks, and maintained at 50mg tid. Low-dose hormone therapy was administered to all patients simultaneously. Patients with fever, infection, or patients with other autoimmune diseases, uncontrolled malignancies, and other chronic diseases were excluded. According to the duration of AZA treatment, 20 patients were divided into 1 month after AZA treatment, 16 patients in 6 months after AZA treatment and 17 patients over 1 year after AZA treatment. Clinical data of patients were collected, including: gender, age, time of first onset, time of medication, clinical symptoms, number of recurrence, EDSS score, serum and cerebrospinal fluid AQP 4-IgG titers, imaging results, rheumatic immunity-related autoimmune antibodies, comorbidities, related adverse drug reactions, etc. Ten healthy volunteers with physical examination in our hospital were selected as the healthy control (HC) group.

The Ethics Committee of the First Affiliated Hospital of Guangxi Medical University approved all of the research procedures, and we obtained informed consent from all of the participants.

### 2.2. Preparation of red blood cells and serum samples

Blood (5ml) was collected from peripheral veins, centrifuged at 3000 rpm for 10 minutes, and 1000ul of upper serum was collected in EP tubes and frozen at -80°C. Centrifugation after normal saline washing, lastly, add 500ul of 40% glycerol solution to 500ul of RBCs at the bottom of the tube, shake it for 1 minute, and freeze it at -80°C. Serum samples from healthy controls were collected and preserved in the same manner.

### 2.3. 6-TGN and 6-MMPN concentrations were determined by HLPC

6-TGN and 6-MMPN was tested by HPLC method published in other studies.

### 2.4. Serum levels of IL-6, IL-18, and NLRP 3 were determined by ELISA

The levels of IL-6, IL-IL-18, and NLRP 3 inflammasome were measured in the enzyme-linked immunoassay kit.

### 2.5. Statistical analysis

SPSS version 25.0 (IBM Corp, Armonk, NY, USA) was used to analyze all of the data. When the measurement data meet the normal distribution, the mean ± standard deviation (x^-^± s) is expressed, independent sample T-test was used for comparisons between two groups, and one-way analysis of variance between multiple groups. Without the normal distribution, data was expressed as median or interquartile spacing, Mann-Whitney U test and Kruskal-Wallis test were used to compare between two or multiple groups. Count data were tested using the Pearson chi-square test or the Fisher exact probability method. Pearson or Spearman analysis of the relationship between continuous variables. *p*< 0.05 was considered to be statistically significant.

## 3. Results

### 3.1. Demographic and clinical features

Myelitis was still the most common symptom in NMOSD patients, followed by optic neuritis, area postrema syndrome (APS), symptomatic narcolepsy or acute diencephalic, and brainstem syndrome. Table 1-1 shows demographic data, disease duration, and clinical symptoms. There were 10 people in the HC group, and the ratio of men to women was 1:4.

**Table 1-1.** Basic clinical characteristics of the NMOSD patients.

|  | 1 month after AZA<br>treatment | 6 months after AZA<br>treatment | over 1 year after AZA<br>treatment |
| --- | --- | --- | --- |
| Patients | 20 | 16 | 17 |
| Male:Female | 1:19 | 1:15 | 1:16 |
| Ages (Y) | 45.05±10.69 | 43.25±11.20 | 42.88±8.91 |
| Optic neuritis | 12 | 10 | 12 |
| Acute myelitis | 17 | 14 | 15 |
| Area postrema<br>syndrome(APS) | 3 | 3 | 1 |
| Acute brainstem<br>syndrome | 1 | 0 | 1 |
| Symptomatic<br>narcolepsy or acute<br>diencephalic | 0 | 1 | 5 |
| Symptomatic cerebral<br>syndrome | 0 | 0 | 0 |
| With AQP4-IgG | 18 | 14 | 9 |
| Total disease<br>duration(M) | 7 (2.43-39.00) | 24 (10.50-66.00) | 46 (25.00-71.00) |

Note: Y=year, M=month.

The mean age was 40.00±16.472 years (Table 1-2).

**Table 1-2.** Comparison of mean age and sex ratio between NMOSD patients and HC.

|  | Case groups | HC | *p* value |
| --- | --- | --- | --- |
| Ages (Y) | 45.05±10.69 | 40.00±16.47 | >0.05 |
|  | 43.25±11.20 | 40.00±16.47 | >0.05 |
|  | 42.88±8.91 | 40.00±16.47 | >0.05 |
| Male: Female | 1:19 1:15 1:16 | 2:8 | >0.05 |

### 3.2. Differences in EDSS scores and ARR before and after medication

The EDSS score is indicated by the median (interquartile range). The median (interquartile range) EDSS scores before and after treatment for each group are shown in Table 2-1. EDSS scores before treatment did not differ statistically significantly between groups (*p*> 0.05). The improvement of EDSS score before and after treatment are shown in Table 2-2. No patients relapsed in the first month after AZA treatment or the six months after AZA treatment, there were two patients who relapsed after more than 1 year of taking AZA (ARR:0 per year). One patient relapsed at two months after taking AZA, another patient relapsed at four years after taking AZA. After relapse, both two patients continued taking AZA, and for more than 1 year without recurrence (ARR before and after treatment, see Table 2-3). The improvement in EDSS score increases and the ARR decreases with an increase in medication time, indicating that long-term AZA treatment improves neurological function and reduces recurrence rates in patients with NMOSD.

**Table 2-1.** Changes in EDSS scores before and after AZA treatment.

| Case groups | EDSS scores |  | *p* value |
| --- | --- | --- | --- |
|  | before AZA treatment | after AZA treatment |  |
| 1 month after treatment | 1.5 (1.0-2.8) | 1.5 (1.0-2.0) | 0.591 |
| 6 months after treatment | 2.0 (1.5-2.4) | 1.0 (1.0-1.5) | 0.005 |
| Over 1 year after treatment | 1.5 (1.3-2.0) | 1.0 (0.0-1.3) | 0.001 |
| All patients | 1.5 (1.0-2.0) | 1.0 (0.5-1.5) | 0.000 |

**Table 2-2.** Changes in EDSS scores before and after AZA treatment.

| Case groups | Patients | Improvement in EDSS score | Kruskal-Wallis test | Comparison among groups |
| --- | --- | --- | --- | --- |
| 1 month after treatment | 20 | 0.0 (0.0-0.5) | H=13.151 | $p1=0.038$ |
| 6 months after treatment | 16 | 0.5 (0.5-1.0) | $p=0.001$ | $p2=1.000$ |
| Over 1 year after treatment | 17 | 1.0 (0.5-1.5) | | $p3=0.002$ |

Note: $p1$ is the comparison result of the 1 month after treatment group and the 6 months after treatment group, $p2$ is the comparison result of the 6 months after treatment group and the over 1 year after treatment, and $p3$ is the comparison result of the 1 month after treatment group and the over 1 year after treatment group.

**Table 2-3.**
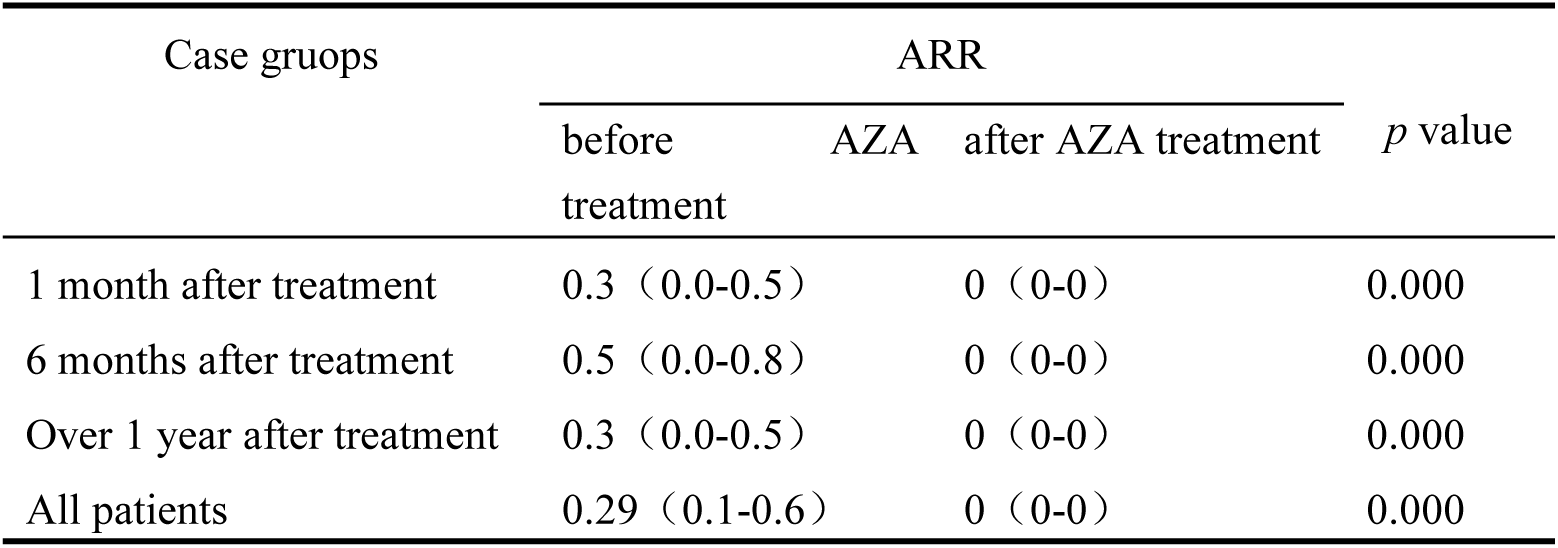
ARR changes in case groups before and after AZA treatment.

### 3.3. The concentrations of 6-TGN and 6-MMPN in NMOSD patients

The mean 6-TGN concentrations in each group were 127.261 ± 39.500pmol/8×10^8^ RBC,171.245±44.261pmol/8×10^8^ RBC,180.815± 67.962pmol/8 × 10^8^ RBC, respectively (Figure 1-1A). EDSS score improvement was positively correlated with the overall 6-TGN concentration (r=0.286, *p*=0.038) as shown in Figure 1-2 A.

**Figure 1-1.**
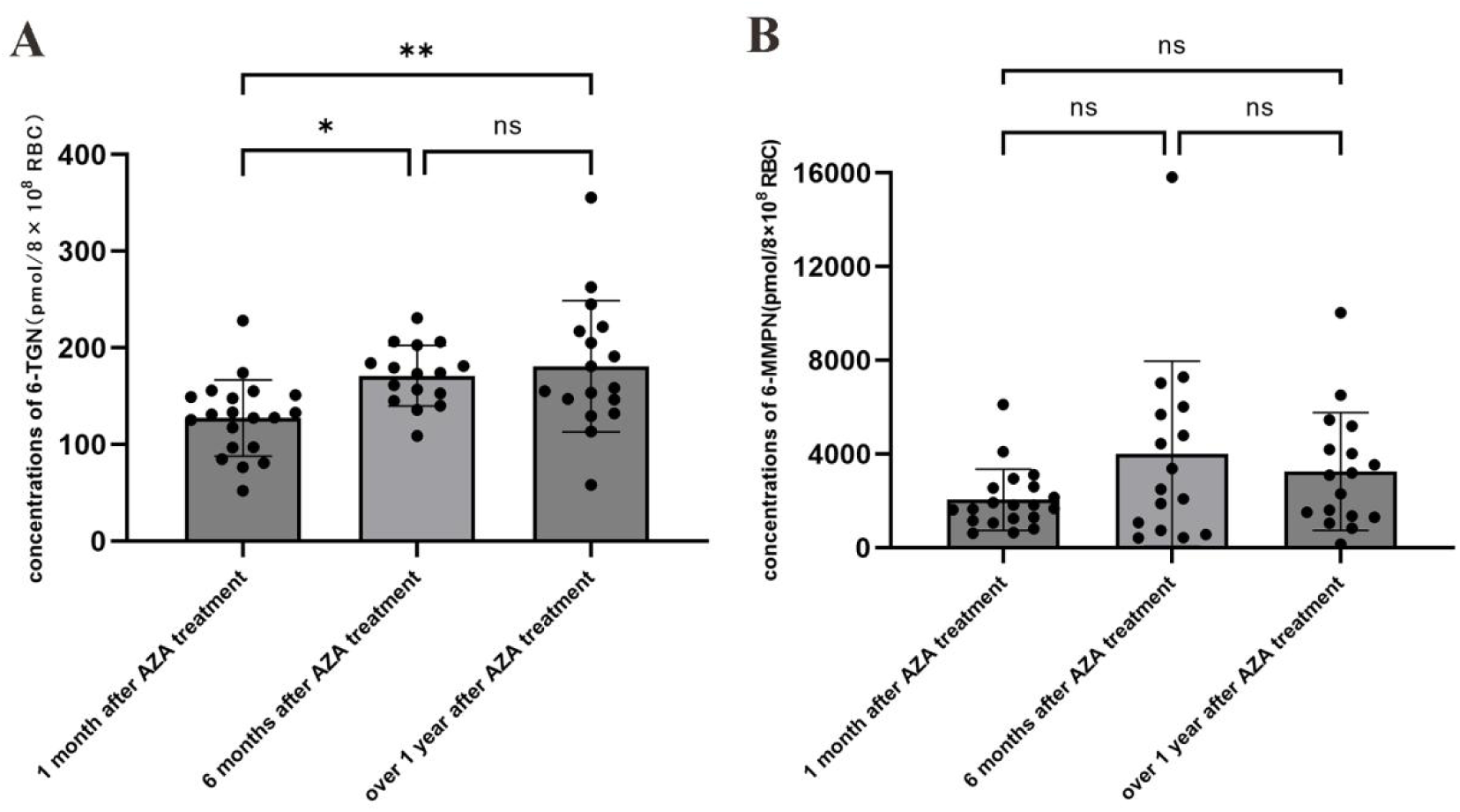
concentrations of 6-TGN (A) and 6-MMPN (B).

The mean concentrations of 6-MMPN were 2048.249 ± 1307.372pmol/8 × 10^8^ RBC,4012.0410 ± 3955.616pmol/8 × 10^8^ RBC, 3257.475 ± 2515.477 pmol /8 × 10^8^ RBC (Figure 1-1B). EDSS score improvement was not correlated with overall 6-MMPN concentrations (r=0.118, *p*=0.401), as shown in Figure 1-2B.

A total of 13 patients (13 / 53) experienced drug-related AEs, of which 2 were treated for one month, 7 for six months, and the rest for over one year. Liver enzyme increase (7 / 53), leukocyte decline (2 / 53), gastrointestinal reaction (1 / 53), hair loss (2 / 53), menstruation decrease (1 / 53) (Table 3-1), were included. The mean of 6-TGN concentrations were 206.503 ± 55.270 pmol /8 × 10^8^ RBC, mean 6-MMPN concentrations were 5862.270 ± 3679.188 pmol / 8×10^8^ RBC. Without AEs in 40 individuals, mean 6-TGN concentrations were 140.188 ± 42.742pmol/8 × 10^8^ RBC, and mean 6-MMPN concentrations were 2108.131 ± 1627.442 pmol / 8×10^8^ RBC. In the group with AEs, the drug concentrations were higher than in the normal group, p=0.000.The ROC area under the curve of 6-TGN concentrations by SPSS25.0 software was 0.871(95%CI:0.770-0.973); the best critical value of 155.604 pmol / 8 × 10^8^ RBC was selected according to the maximum Youden index, resulting in the maximum Youden index value of 0.673 with corresponding sensitivity of 92.3% and specificity of 75%. The ROC area under the curve of 6-MMPN concentrations was 0.898 (95%CI:0.816-0.980); the best cut-off value of 2583.168 pmol / 8 × 10^8^ RBC was selected according to the Youden index at its maximum, yielding a maximum Youden index value of 0.775, corresponding sensitivity of 100% and specificity of 77.5% (Figure 2).

**Table 3-1.** Drug-related AEs

| AEs | Patients (%) |
| --- | --- |
| Elevated liver enzymes | 7 (13.2%) |
| White blood cell decline | 2 (3.8%) |
| Gastrointestinal reaction | 1 (1.9%) |
| lipso-trichia | 2 (3.8%) |
| relative amenorrhea | 1 (1.9%) |

**Figure 1-2.**
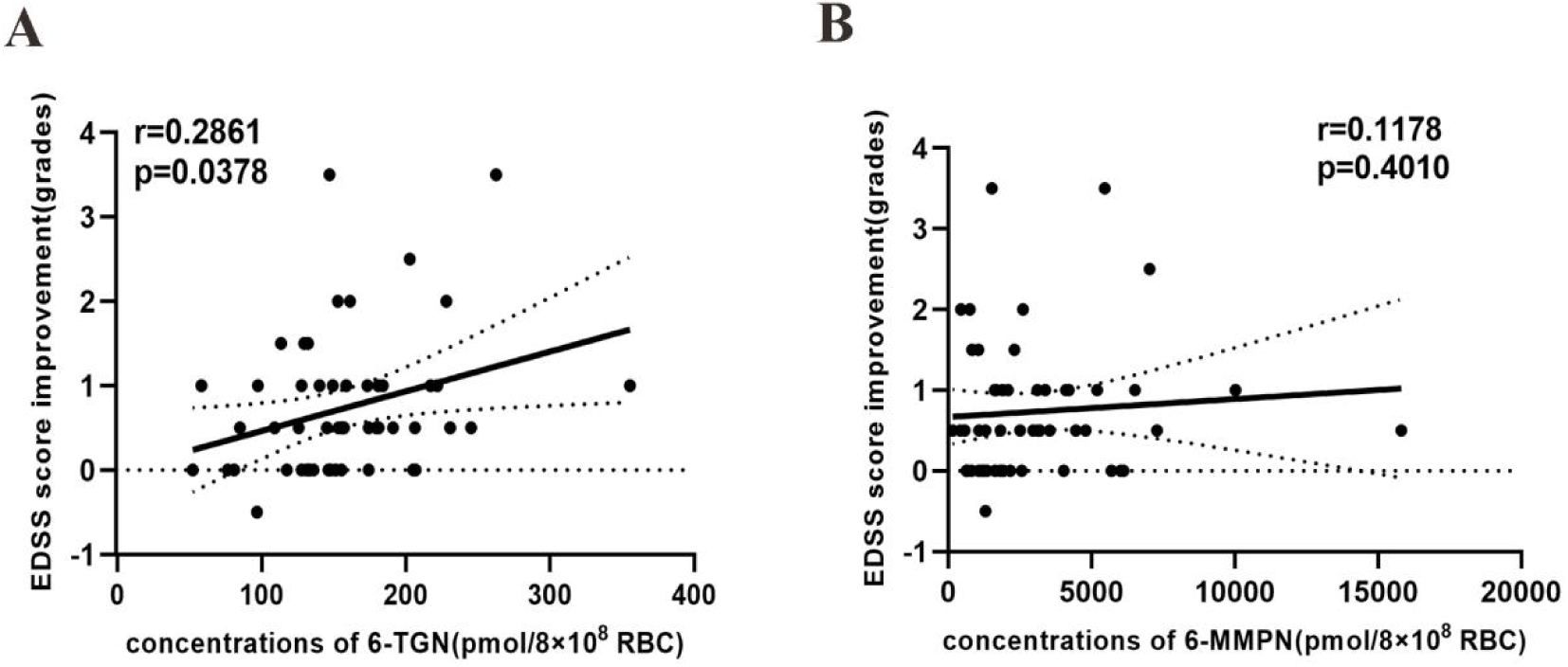
Relationship between 6-TGN(A)/6-MMPN(B) concentrations and EDSS score improvement.

**Figure 2.**
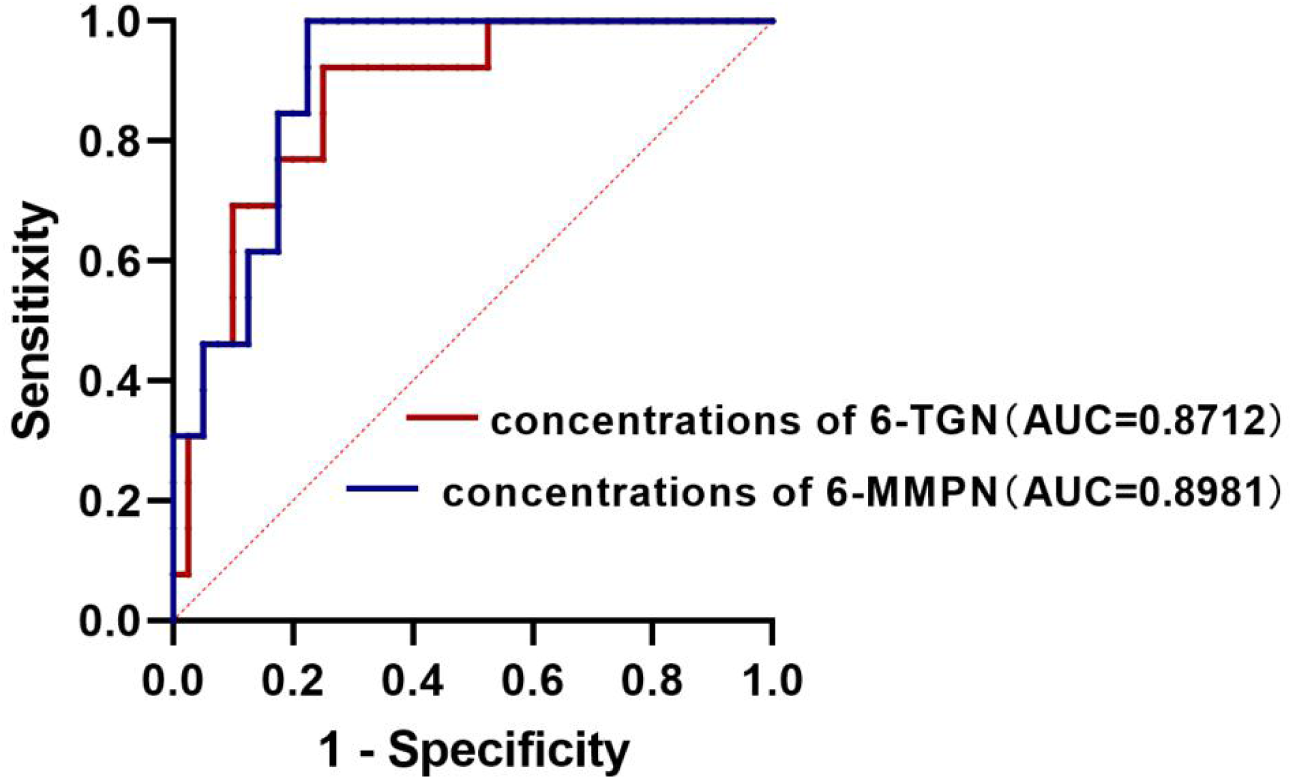
ROC curves of RBC 6-TGN and 6-MMPN concentrations as diagnostic biomarkers.When 155.604 pmol / 8×10^8^ RBC was selected as the cut-off value, 6-TGN as a diagnostic biomarker for the occurrence of adverse drug reactions had a sensitivity of 92.3%, specificity of 75%.When 2583.168pmol/8×10^8^ RBC was selected as a cut-off value, 6-MMPN as a diagnostic biomarker for the occurrence of adverse drug reactions had a 100%, specificity of 77.5%.

### 3.4. Serum levels of IL-6, NLRP3 and IL-18 in NMOSD patients and HC

The IL-6 levels in the case groups were 8.08 ± 1.38pg/ml, 8.06 ± 1.61pg/ml, 7.81±1.98pg/ml, respectively. The IL-6 levels in the HC were 6.15 ± 1.15pg/ml, lower than the case groups (Fig3-1A). The NLRP3 inflammasome levels in the case group were respectively 259.18 ± 34.73pg/ml, 281.65 ± 40.35pg/ml, 274.63 ± 44.29pg/ml; NLRP3 inflammatory factor levels in HC were 203.00±17.64pg/ml, lower than the case group, *p*<0.05 (Fig3-1B). The IL-18 levels in the case groups were 40.25 ± 8.83pg/ml, 44.37 ± 13.70pg/ml, 41.94 ± 10.99pg/ml; the IL-18 levels in HC were 34.11±9.17pg/ml, in comparison with the case groups, it was lower, but not statistically significant (Fig3-1C). 6-TGN concentrations did not correlate with IL-6, NLRP3 inflammasome or IL-18 levels. The levels of NLRP3 inflammasome and IL-18 were associated with the levels of IL-6 (r=0.360, *p*=0.008; r=0.477, *p* <0.001); Levels of IL-18 did not appear to be associated with NLRP3 inflammasome activity (Figure 3-2A, B, C). The levels of NLRP3 inflammasome, IL-18, and IL-6 did not correlate with EDSS scores.

**Figure 3-1.**
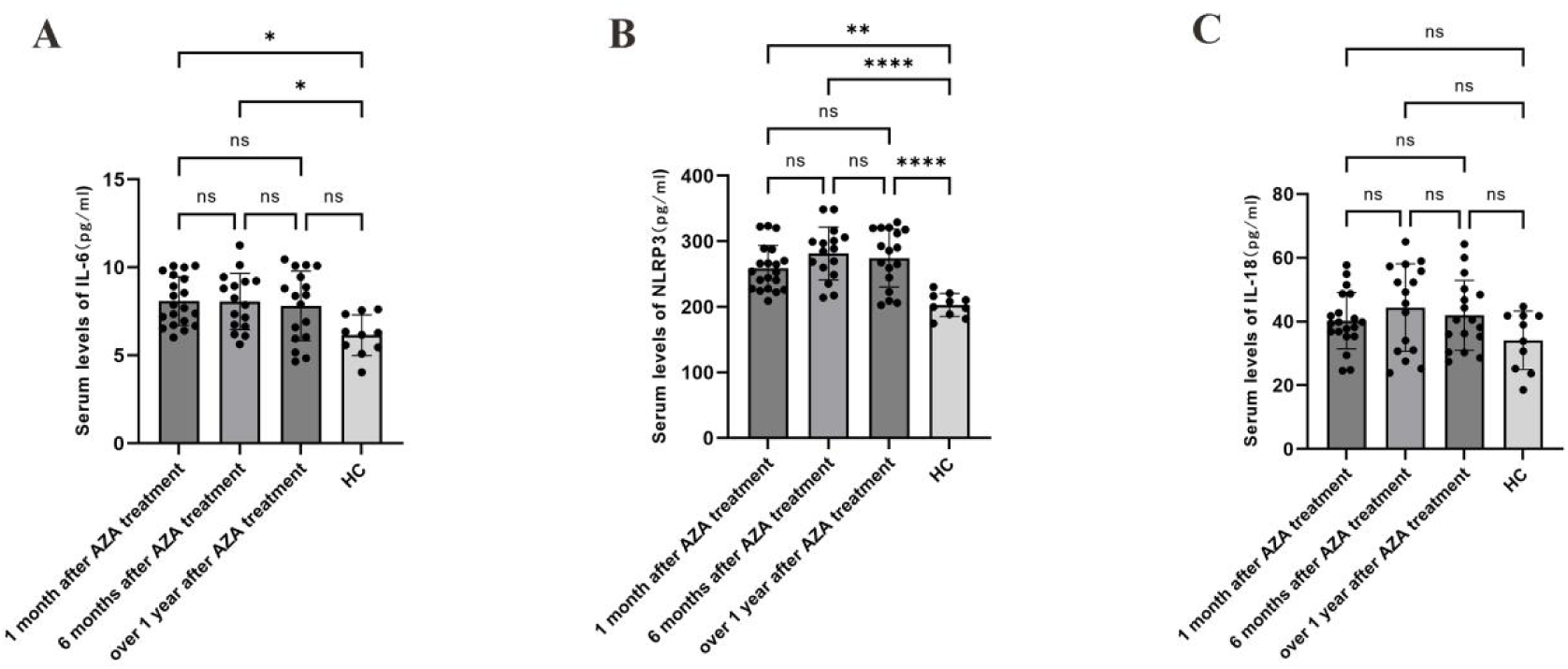
IL-6 (A), NLRP 3 inflammasome (B), IL-18 (C) levels in case groups and healthy controls.

**Figure 3-2.**
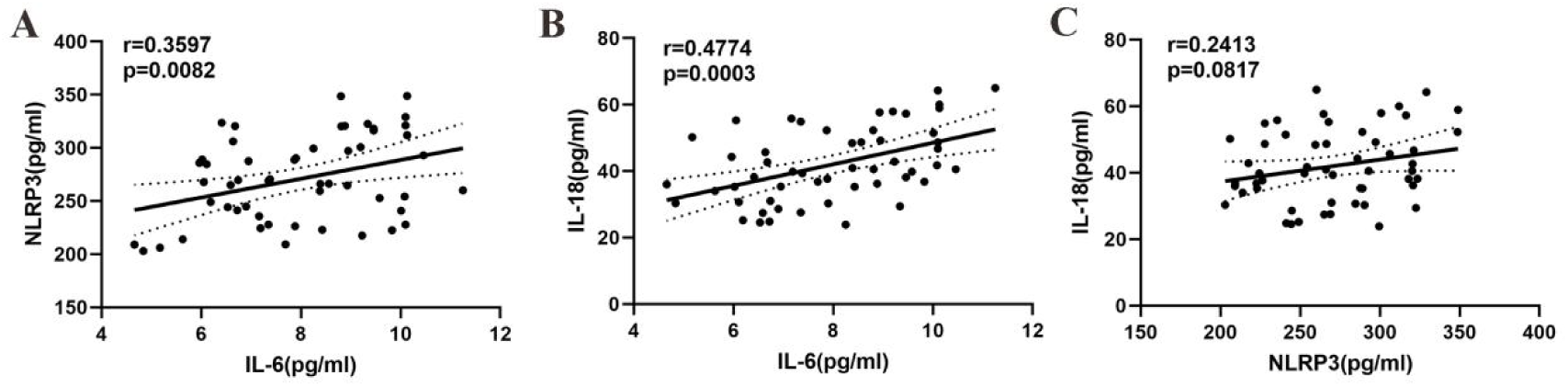
Association between the NLRP3 inflammasome and IL-6 (A), IL-18 and IL-6 (B), and IL-18 and the NLRP3 inflammasome (C).

### 3.5. Correlation between cytokines and clinical features

22 NMOSD patients had associated rheumatic immune-related antibodies, including anti-nuclear antibodies, anti-SS-A antibodies, anti-keratin antibodies and anti-Ro-52 antibodies, but no patients met the diagnostic criteria for related diseases. Cytokine levels were not significantly different between patients with and without rheumatic immune antibodies (*p*> 0.05) in this study. Of all patients, 19 with painful spasm or nocicsensitivity, mean NLRP3 levels were 286.932 ± 40.238pg/ml, 34 without pain, mean NLRP3 levels were 261.946 ± 37.666 pg/ml (*p*<0.05, Figure 4).

**Figure 4.**
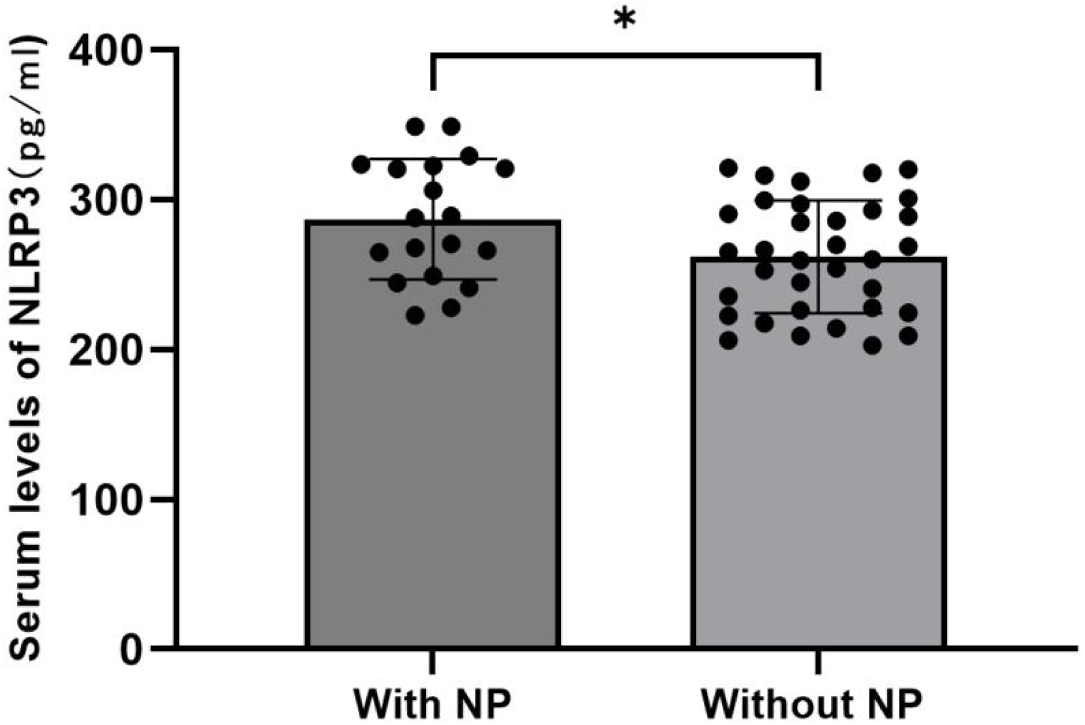
Comparison of NLRP 3 inflammasome levels in NMOSD patients with or without NP.

## 4. Discussion

In this study, all patients experienced 45 relapses before and after medication, with a maximum EDSS score of 8 during follow-up. Based on the results of this study, ARR decreased significantly and EDSS score improved significantly after AZA treatment, suggesting that AZA was an effective drug. AZA is rapidly converted to 6-mercaptopurine in vivo and further to thiopurine nucleotides by a series of competitive enzymes, including 6-TGN and 6-MMPN. Immunotherapeutic effects of 6-TGN and 6 MMPN are achieved by inhibiting nucleic acid biosynthesis and preventing lymphocyte proliferation[^15^]. According to the results of this study, the concentrations of 6-TGN gradually increased, and the concentrations remained relatively stable for about six months, EDSS scores improved positively with 6-TGN concentrations, indicating that as 6-TGN accumulates in vivo, the immunosuppressive effects of the drug begin to be felt. 6-MMPN concentrations did not differ significantly between groups, and there was no correlation between concentrations and improvement in EDSS scores. According to a previous study, patients with NMOSD who were not relapsed had significantly higher 6-TGN and 6-MMPN concentrations than patients who were relapsed, and red blood cell concentrations of 6-TGN (r=0.679, *p*=0.0001) and 6-MMPN (r=0.493, *p*=0.004) were associated with improvements in ARR. 6-MMPN is a strong inhibitor of de novo purine synthesis that blocks proliferation in various types of lymphoid cell lines^[16]^. This study did not include patients with acute recurrences, so it is unclear whether 6-MMPN affects the course of NMOSD patients. According to the current literature review, 6-MMPN concentration is associated with acute recurrence but not severity of disease.

Before receiving AZA, all patients were tested for TPMT activity. Some patients, however, developed AZA-related adverse effects, such as liver enzyme elevation, leukocyte decline, and alopecia, which may be due to excess drug concentrations. The concentrations of 6-TGN and 6-MMPN in the AEs group were higher than those in the normal group (*p*=0.000). The increased risk of AEs was calculated when the 6-TGN concentration reached 155.604 pmol /8×10^8^ RBC or 6-MMPN reached 2583.168 pmol / 8×10^8^ RBC. Our previous study examined whether 6-TGN concentrations differed in leukocyte decreased patients, and the results showed that 6-TGN concentrations increased^[17]^. Patients with high drug concentrations should have their dosage adjusted in time, reducing the likelihood of drug withdrawal and AEs.

IL-6 could further induce AQP 4-IgG production in B cells and promote the pathophysiology of NMOSD^[18]^. Previous studies suggested that IL-6 levels increased in CSF and serum of NMOSD patients ^[19]^. IL-6 receptor antagonists had been used in the treatment of NMOSD. In this study, serum IL-6 levels gradually decreased with treatment duration, the serum IL-6 levels of NMOSD patients in the 1 month and 6 month after AZA treatment groups were higher than those of HC. It appears that IL-6 plays a proinflammatory role in NMOSD, but it may not be associated with disease severity. IL-6 levels decrease with increasing medication time, which can reduce disease relapse. However, there is no direct relationship between IL-6 and 6-TGN concentrations. During a two-year follow-up period, serum IL-6 levels were associated with clinical relapse risk and disease severity in another Brazilian study ^[20]^. The conclusions of the two studies are not contradictory since this study did not include patients with acute relapse.

As a result of this study, NLRP3 levels were higher in patients than in HC, and its levels were correlated with IL-6 levels, suggesting that NLRP 3 plays a role in the inflammatory response process of NMOSD. In this study, there was no correlation between NLRP 3 levels and 6-TGN concentration or EDSS score, so it is not possible to determine the effect of AZA on NLRP3, and in NMOSD remission, serum NLRP3 levels may not be related to severity. NLRP 3 inflammasome and mtDNA levels were significantly higher in CSF of NMOSD and MS patients, according to Yu Peng et al, and CSF NLRP3 levels in NMOSD patients were significantly associated with EDSS score, mtDNA, IL-1 β, IL-6 and IL-17, suggesting that NLRP 3 inflammasome-mediated cytopyroptosis after mitochondrial injury may play an important role in the pathogenesis of inflammatory demyelinating disease^[9]^. According to our study and Yu Peng et al., NMOSD patients have significantly higher NLRP 3 inflammasomes than normal individuals, and NLRP 3 is associated with IL-6. According to our study, there is no correlation between NLRP 3 inflammasome level and EDSS score. NLRP 3 inflammasome mediates caspase-1 activation and secretion of pro-inflammatory cytokines IL-1 β, IL-18, whose abnormal activation is associated with inflammatory diseases, mainly promoting the activation of inflammatory factors and inducing pyroptosis^[8]^. Yu Peng et al found that CSF NLRP3 levels were significantly associated with IL-1 β. However, serum NILP 3 levels were not correlated with levels of IL-18 in this study. Studies have shown that IL-1 β can induce the migration of helper T cells to the CNS, which releases IL-6 and IL-17, and triggers an inflammatory cascade that damages the CNS in NMOSD^[21]^.

Early studies showed that IL-18 could enhance the production of including IL-4, IL-5, IL-13, GM-CSF and TNF^[22]^; IL-18 could promote the expression of perforin perforin and FASL, and enhance the cytotoxic of NK cells^[23]^. The finding that IL-18 levels are increased after erythrin-induced excitatory brain injury and hypoxic-ischemic brain injury suggests that IL-18 plays a role in CNS and causes cell damage^[24]^. Patients with MS, Alzheimer’s disease, epilepsy, myasthenia gravis and Gililan-Barre syndrome have elevated serum IL-18 levels^[25]^. Patients’ serum IL-18 levels were correlated with IL-6 levels, and higher than controls, but there was no statistically significant difference between case groups and HC groups. When NMOSD patients are in remission, there is no correlation between IL-18 and 6-TGN concentration, EDSS score, or NLRP3 levels. IL-18 was secreted by NLRP 3 inflammasome, but there was no obvious correlation, and further research is needed to determine the mechanism.

Sensory system involvement is common in patients with NMOSD, such as hypoesthesia, hypersensitivity, numbness, and pain. Painful spasticity and limb pain symptoms in NMOSD fit the NP definition^[26]^. NP is associated with neuronal sensitization in the nervous system. In addition to neurons, NP also involves the of immune cells, glial cells, etc^[27]^. In this study, 19 patients remained with painful spasms or pain sensitivity during blood collection, and NLRP 3 levels in serum were higher than in patients without NP. It appears that NLRP3 inflammasome is also involved in the pathophysiology of NP.

21 patients had serum antibodies, such as anti-SS-A antibodies and anti-Ro-52 antibodies. The serum levels of IL-6, NLRP 3, and IL-18 did not differ with or without these rheumatic immune antibodies, suggesting that NMOSD patients may showed a small effect of rheumatic immune antibodies on their immune inflammatory status.

## 5. Conclusion

As 6-TGN concentration increased, patients treated with AZA showed improved neurological function and decreased ARR. AZA-related AEs can occur when 6-TGN concentrations reached 155.604 pmol/8×10^8^ RBC or 6-MMPN concentrations reached 2583.168 pmol/8×10^8^ RBC. The serum levels of IL-6, NLRP3, and IL-18 were higher than those of HC during the remission period of NMOSD, suggesting their involvement in the development and development of NMOSD. An excess of NLRP3 is associated with NP in NMOSD patients.

## Data Availability

All data produced in the present study are available upon reasonable request to the authors
All data produced in the present work are contained in the manuscript
All data produced are available online at

## Acknowledgment

This work was supported by the the Guangxi Health Committee of China (S2019093). The founding source did not play any other role than financial support. The authors declare no competing interests.

